# Estimation of introduction and transmission rates of SARS-CoV-2 in a prospective household study

**DOI:** 10.1101/2023.06.02.23290879

**Authors:** Michiel van Boven, Christiaan H. van Dorp, Ilse Westerhof, Vincent Jaddoe, Valerie Heuvelman, Liesbeth Duijts, Elandri Fourie, Judith Sluiter-Post, Marlies A. van Houten, Paul Badoux, Sjoerd Euser, Bjorn Herpers, Dirk Eggink, Marieke de Hoog, Trisja Boom, Joanne Wildenbeest, Louis Bont, Ganna Rozhnova, Marc J. Bonten, Mirjam E. Kretzschmar, Patricia Bruijning-Verhagen

**Affiliations:** Julius Center for Health Sciences and Primary Care, University Medical Center Utrecht, Utrecht University, Utrecht, the Netherlands; Center for Complex Systems Studies (CCSS), Utrecht University, Utrecht, The Netherlands; Department of Pathology & Cell Biology, Columbia University Irving Medical Center, New York, United States; Erasmus Medical Center, Rotterdam, the Netherlands; Spaarne Gasthuis, Hoofddorp, the Netherlands; Regional Public Health Laboratory Kennemerland, Haarlem, the Netherlands; National Institute for Public Health and the Environment, Bilthoven, the Netherlands; Department of Paediatric Infectious Diseases and Immunology, Wilhelmina Children’s hospital, University Medical Center Utrecht, the Netherlands; BioISI—Biosystems & Integrative Sciences Institute, Faculdade de Ciências, Universidade de Lisboa, Lisbon, Portugal

## Abstract

Household studies provide an efficient means to study transmission of infectious diseases, enabling estimation of individual susceptibility and infectivity. A main inclusion criterion in such studies is often the presence of an infected person. This precludes estimation of the hazards of pathogen introduction into the household. Here we use data from a prospective household-based study to estimate SARS-CoV-2 age- and time-dependent household introduction hazards together with within household transmission rates in the Netherlands from August 2020 to August 2021. Introduction hazards and within-household transmission rates are estimated with penalized splines and stochastic epidemic models, respectively. The estimated hazard of introduction of SARS-CoV-2 in the households was lower for children (0-12 years) than for adults (relative hazard: 0.62; 95%CrI: 0.34-1.0). Estimated introduction hazards peaked in mid October 2020, mid December 2020, and mid April 2021, preceding peaks in hospital admissions by 1-2 weeks. The best fitting transmission models include increased infectivity of children relative to adults and adolescents, such that the estimated child-to-child transmission probability (0.62; 95%CrI: 0.40-0.81) was considerably higher than the adult-to-adult transmission probability (0.12; 95%CrI: 0.057-0.19). Scenario analyses show that vaccination of adults could have strongly reduced infection attack rates in households and that adding adolescent vaccination would have offered limited added benefit.

## Introduction

Transmission of SARS-CoV-2 occurs predominantly in indoor settings such as public transportation, workplaces, schools, and households [1–4]. Infection can cause the respiratory and systemic disease COVID-19, but in general severity and progression of the disease are mild [5, 6]. This has hampered, even despite huge research efforts, to accurately quantify variations in susceptibility and infectiousness, and how these depend on host characteristics such as age and sex, type of infecting strain, pre-existing immunity, and vaccination.

Household studies are considered the gold standard for the study of infectious disease transmission, as they provide a setting in which transmission events can be pinned down to one or a small number of potential infectors [7–18]. Classical analyses of household data use statistical regression techniques to estimate the fraction of persons that are infected over the course of a household outbreak (the secondary attack rate or SAR), stratified by person-type and household characteristics [19]. For SARS-CoV-2, a meta analysis of 54 studies has revealed that secondary attack rates are higher when the index case is symptomatically infected, that transmission to adults occurs more often than to children, that transmission to spouses occurs more often than to other family contacts, but that there are no significant sex-differences in attack rates [4]. These studies, while providing valuable information, do not provide estimates of parameters that have a biological interpretation, and in particular do not provide insight in the rates of direct person-to-person transmission. As a consequence, they also do not lend themselves to extrapolation and scenario analyses. In addition, most studies use a reactive design in which households are included only after an infected person has been detected in the household (see [20] for an exception). This makes these studies vulnerable to bias, e.g., household-size biased inclusion and bias toward inclusion of households with more severely infected index cases. Also, as households are only included after the first infection these studies cannot estimate the rates at which infections are introduced into the household.

We propose the prospective household-based cohort study as an attractive crossover of the reactive household study and the prospective cohort study. While classical person-based prospective cohort studies in principle can provide high-quality information on risk factors and confounding variables, they are also inefficient if the outcome (infection) occurs infrequently [21]. This inefficiency is remedied by employing a household based inclusion, as it increases the number of events (i.e. infections). We illustrate this by using data from a prospective household study carried out in the Netherlands in the first year of the SARS-CoV-2 pandemic. The study contains a total of 1, 209 persons distributed over 307 households, of which 59 are infected during the study period. We analyze the data in a Bayesian framework using survival analysis for estimation of the hazards of introduction of SARS-CoV-2 into households, and stochastic SEIR epidemic models for estimation of within household transmission rates. As it is known that contacts between household members do not occur randomly [22], we stratify the analyses by person-type (child, adolescent, adult), and select the most likely contact structure based on statistical evaluation of competing models. We show that precise estimates of the type-specific introduction rates can be obtained together with the person-to-person transmission rates, and explore the impact of different vaccination strategies on reducing (1) the role of households as a multiplier of infection and (2) the probabilities of infection of specific persons (e.g., adults).

## Results

### Introduction of SARS-CoV-2 into households

Inclusion of households has been fairly uniform from September 2020 until January 2021, and decreasing from January 2021 onward (Fig. 1). Most households are included for the maximal follow-up period of 161 days, and SARS-CoV-2 has been introduced and established in almost one out of five households (59 of 307, 19%).

**Figure 1:**
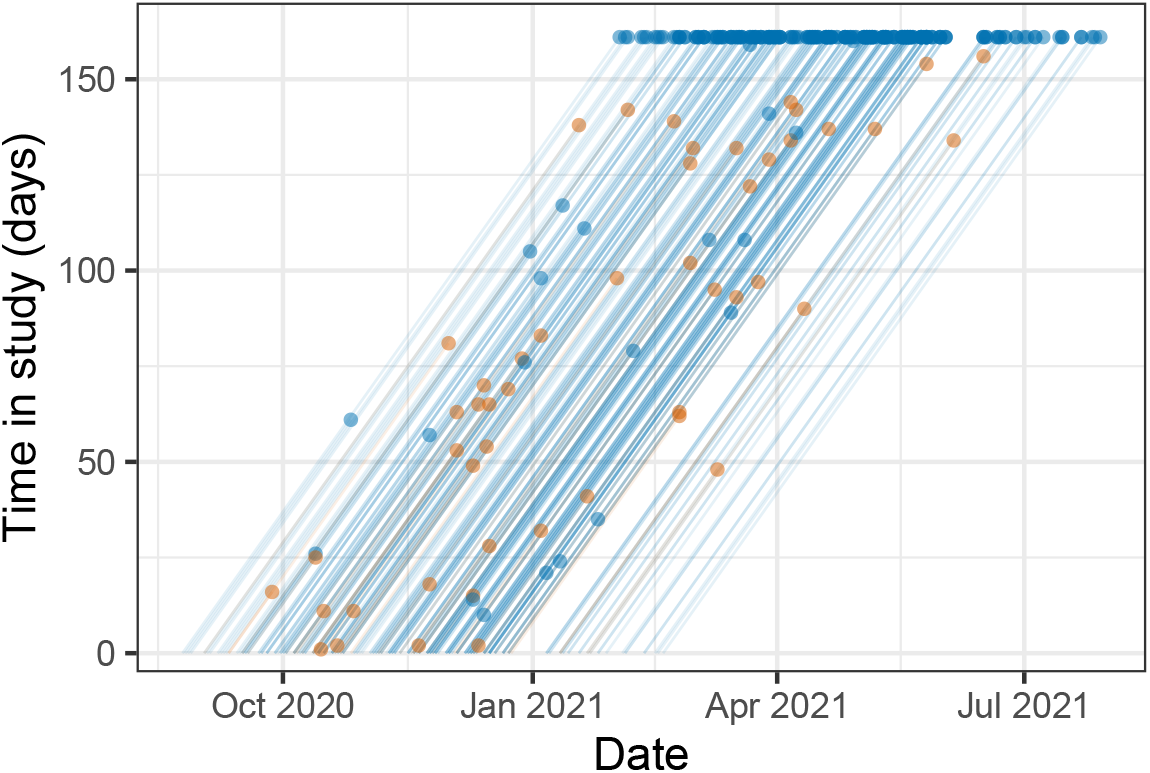
Lexis diagram of the study population. Lines show households from the date of inclusion in the study to infection of the first household member (brown dots), completion of the inclusion period without infection (blue dots at time in study of 161 days) or dropout (blue dots with time in study shorter than 161 days). Date of inclusion of the first household was 24 August 2020, and last date of the study was 29 July 2021.

Figure 2 shows the per-person hazard of introduction into the household for adults together with the 7-day smoothed hospital admissions in the Netherlands. It illustrates that per-person introduction hazards range from approximately 0.0002 per adult per day in February 2021 and July 2021 to almost 0.001 per adult per day in December 2020. Specifically, there are three peaks in the introduction hazards, viz. 0.00080 (95%CrI: 0.00039*−*0.0015) per adult per day in mid-October 2020, 0.0010 (95%CrI: 0.00058*−* 0.0017) in mid-December 2020, and 0.00074 (95%CrI: 0.00042 *−* 0.0014) in early April 2021. These peaks consistently precede peaks in the number of hospital admissions with SARS-CoV-2 by 1 to 2 weeks. The per-person hazards of introduction of SARS-CoV-2 for children and adolescents are estimated relative to adults, and indicate that children (relative hazard 0.62, 95%CrI: 0.34 *−* 1.0) have a lower introduction rate into the household than adults, while the introduction hazard of adolescents is similar to that of adults (relative hazard 0.97, 95%CrI: 0.52 *−* 1.7).

**Figure 2:**
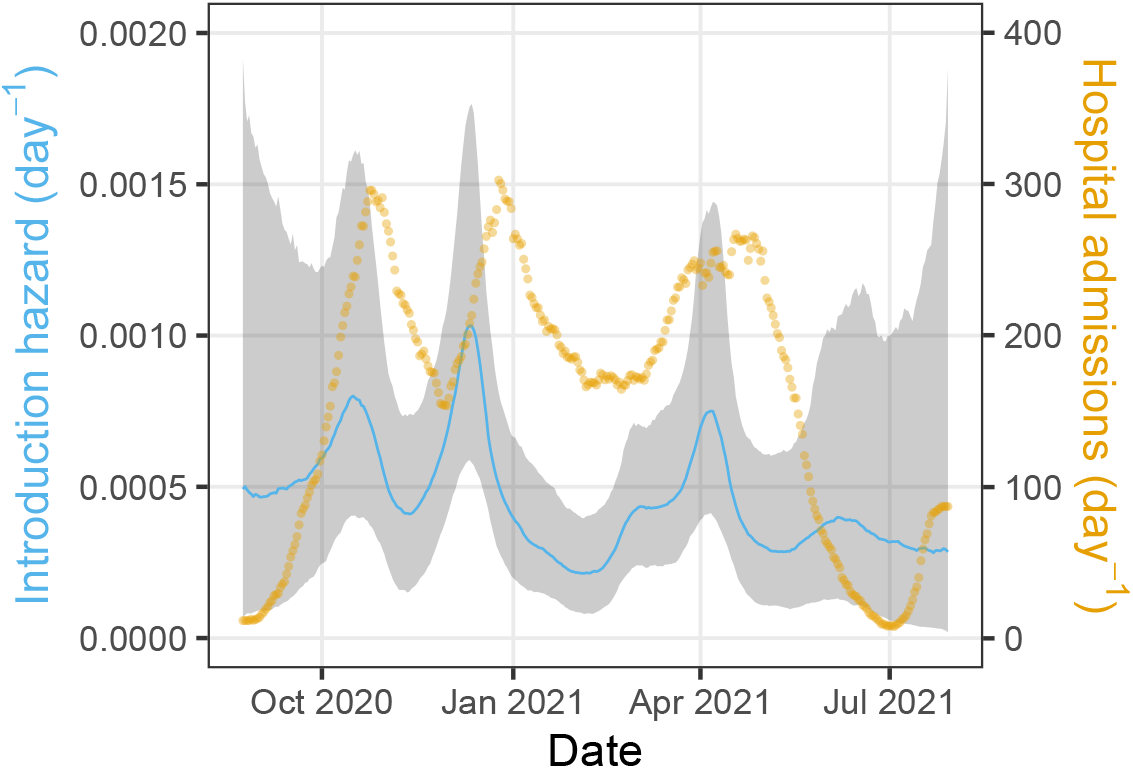
Estimates of the household introduction hazard for adults. Shown are the posterior median of the introduction hazard per person (blue line) with associated 95% credible envelope (gray area). Household introduction hazards of children and adolescents are obtained by multiplication of the hazard for adults with the relative introduction hazards for children and adolescents (Table 2). Also presented are the daily number of hospitalisations (yellow dots). To remove weekday effects the number of hospitalisations are represented by a 7-day moving average centered around the current day.

To explore whether the hazards of introduction can be explained by simpler parametric functions with small number of parameters we perform a number of sensitivity analyses. For instance, a scenario which assumes fixed person-specific introduction hazards yield estimates of the introduction hazards of 0.00030 (0.00018 *−* 0.00047) per day for children, 0.00053 (0.00031 *−* 0.00086) per day for adolescents, and 0.00052 (0.00039 *−* 0.00067) per day for adults. Hence, this model confirms that the introduction hazard is lower for children than for adolescents and adults. However, this fixed-hazard model has low statistical support compared to the spline model (ΔLOO_IC > 5 and ΔWBIC > 10 in favor of the spline model). Similarly, low-order polynomial extensions of this model also have low statistical support (not shown).

### Within-household transmission of SARS-CoV-2

A total of 59 out of 307 households had a detected SARS-CoV-2 introduction over the course of the study, and in these 59 households 119 of 237 persons had documented SARS-CoV-2 infections. Of these 119 infections, 77 are considered primary or co-primary cases that introduced the infection in the households. Total numbers of children, adolescents, and adults in these households are 89, 31, and 117, corresponding numbers of primary and co-primary cases are 21, 8, and 48, and corresponding numbers of household infections are 19, 3, and 20.

The household data are analyzed with a suite of transmission models that vary in their assumptions on the person-to-person transmission rates. An overview of the scenarios is given in Table 1. The analyses show that models that do not stratify the population by person-type (‘no stratification’), or models that assume a separate transmission parameter for each person-type combination (‘full model’) perform less well than models of intermediate complexity. In particular, the ‘variable infectivity’ model with an additional parameter for child-to-child transmission performs best both with regard to LOO IC and WBIC.

**Table 1:** Comparison of household models using information criteria. Model selection is based on the information criteria LOO_IC and WBIC. Shown are results for models that do not stratify the population by age (‘no stratification’), and that stratify the population into children (0-12 years), adolescents (12-18 years), and adults (over 18 years). Stratified transmission models are considered in which susceptibility of different age groups is estimated while infectivity is assumed to be identical for different age group (‘variable susceptibility’), in which infectivity is estimated while susceptibility is fixed (‘variable infectivity’), and in which both susceptibility and infectivity are estimated (‘proportionate mixing’). Within the ‘variable infectivity’ model we further consider sub-models with transmission rates for child-to-child, adolescent-to-adolescent, and adult-to-adult transmission. A saturated model with a separate parameter for each transmission rate is also considered (‘full model’). The number of within-household parameters is indicated by *n*. All models assume density-dependent transmission.

| Model | n | LOO_IC | WBIC |
| --- | --- | --- | --- |
| No stratification | 1 | 143.9 | 141.7 |
| Variable susceptibility | 3 | 145.4 | 140.1 |
| Variable infectivity | 3 | 138.1 | 133.8 |
| with child-to-child transmission | 4 | 135.1 | 129.0 |
| with adolescent-to-adolescent transmission | 4 | 140.5 | 135.1 |
| with adult-to-adult transmission | 4 | 139.4 | 133.7 |
| Proportionate mixing | 5 | 142.0 | 134.1 |
| Full model | 9 | 142.6 | 130.9 |

Parameter estimates of the preferred model indicate that children are more infectious overall than adults and adolescents (relative infectivity: 2.4, 95%CrI: 1.4 *−* 5.9), and that child-to-child transmission has the highest estimated transmission rate (0.97 per infectious period, 95%CrI: 0.51−1.7)(Table 2). Using the estimates of Table 2, Fig. 3 presents the estimated probabilities of person-to-person transmission. Here the highest estimated transmission probability is from child-to-child (0.62, 95%CrI: 0.40 − 0.81), followed by transmission from child-to-adult and child-to-adolescent (0.25, 95%CrI: 0.13 − 0.40), and from adult to all other person-types (0.12, 95%CrI: 0.057 − 0.19). All other transmission routes have lower estimated transmission probabilities (≤ 0.10).

**Table 2:** Estimates of within-household transmission parameters. Parameter estimates are shown for the variable infectivity model with separate child-to-child transmission (cf. Table 1). Estimates are represented by posterior medians and 2.5% and 97.5% posterior quantiles, and are based on 1,000 samples from the posterior distribution. Notice that the introduction hazard parameters of children and adolescents are relative to the introduction hazard in adults (cf. Fig. 2).

| Parameter | Estimate | 95%CrI |
| --- | --- | --- |
| Transmission rate among adults<br>(per person per infectious period) | 0.13 | (0.059-0.22) |
| Infectiousness of children relative to adults | 2.4 | (1.4-5.9) |
| Infectiousness of adolescents relative to adults | 0.85 | (0.076-3.9) |
| Transmission rate among children<br>(per person per infectious period) | 0.97 | (0.51-1.7) |
| Relative introduction hazard for children | 0.62 | (0.34-1.0) |
| Relative introduction hazard for adolescents | 0.97 | (0.52-1.7) |

**Figure 3:**
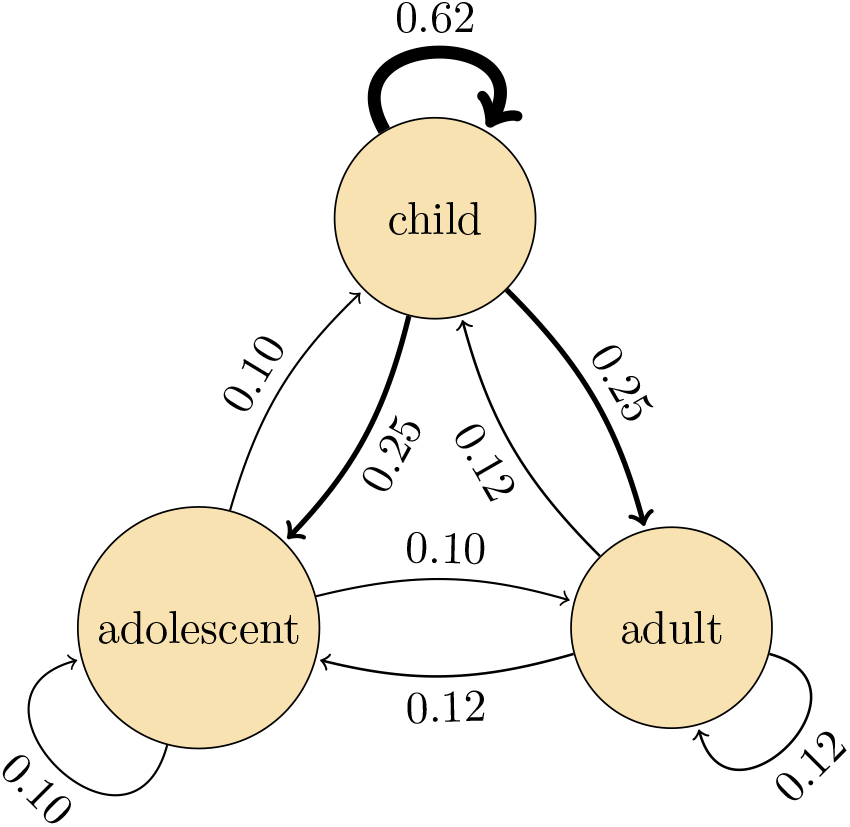
Estimated person-to-person transmission probabilities. Shown are posterior medians of the infectious contact probabilities, i.e. the probabilities that a transmission event would have occurred from an infected person over its infectious period if the contacted person had not already been infected by another person. Infectious contact probabilities are calculated from the person-to-person transmission rates per infectious period *β*_*ij*_ (Table 2) and the Laplace transform of the scaled infectious period distribution: 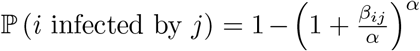, where *α* = 50 is the shape parameter of the infectious period probability distribution.

Sensitivity analyses are performed with respect to the distribution of the infectious period and assumptions on the primary case(s) in the household. As final size distributions are invariant with respect to the mean of the infectious period [23], we focus on how the results are affected by variation in the infectious period distribution. We consider scenarios with an infectious period of fixed duration, and one with an infectious period with more variation than in the default scenario. The parameter estimates of the scenario with fixed infectious period are very close to those reported in Table 2 (not shown). The same is true if variation in the infectious period is increased such that the 95% coverage is 0.61 − 1.48 (gamma shape and rate parameter both equal to 20) instead of 0.74 − 1.30 in the default scenario. Second, we consider a scenario in which we assume that all cases that had originally be designated as co-primary cases (i.e. infected from outside of the household) had actually been infected within the household [3]. In this scenario, the precision with which introduction hazards can be estimated is decreased, while precision of within-household transmission parameter estimates is increased. Noteworthy, the overall within-household transmission rate between adults increases, while the three peaks of the introduction hazard are still noticeable but are decreased in size. Overall patterns of within-household transmission, including the high estimated child-to-child transmission rates, remain as in the main analyses.

### The impact of vaccination on household transmission

The above preparations enable quantification of the role of households as a multiplier of infection. We focus on the secondary attack rate (SAR) and the probability that a focal adult in the household is infected. Table 3 shows the results for various household compositions and vaccination scenarios. Specifically, we consider two vaccination scenarios, one in which all adults in the household are vaccinated, and another one in which both adolescents and adults are vaccinated. In both cases we assume a leaky vaccine that is highly effective in preventing infection (VE_*S*_ = 0.9) but does not reduce infectiousness (VE_*I*_ = 0). Due to the high estimated infectiousness of children, estimated SARs are invariably higher if a child is the primary case rather than an adolescent or adult. Differences in outbreak size can be substantial, especially in larger households. For instance, in households of six persons the estimated SAR is 0.50 if the child is the primary case, but less than 0.25 if it is the adolescent or adult. Noteworthy, estimates are less precise in households with one or more adolescents due to the relatively small number of adolescents in our study.

**Table 3:** Estimated secondary attack rates without and with vaccination. Shown are inferred secondary attack rates (SARs) for various household compositions. Estimates are represented by posterior medians and 2.5% and 97.5% posterior quantiles. Households consist of either a child and an adult (rows 1-2), an adolescent and an adult (rows 3-4), two children and two adults (rows 5-6), two adolescents and two adults (rows 7-8), or two children, two adolescents, and two adults (rows 9-11), thus including the most common household compositions in the Netherlands (rows 1-8). For each household composition, SARs are calculated for all possible primary cases. Vaccination scenarios are considered in which adults are vaccinated, or in which both adults and adolescents are vaccinated. The vaccine is assumed to be 90% effective (VE_*S*_ = 0.9) in preventing infection. NA: households do not contain an adolescent. Estimates are based on 1,000 samples from the posterior distribution.

| Household composition (numbers) |  |  | Primary case | Vaccination scenario |  |  |  |  |  |
| --- | --- | --- | --- | --- | --- | --- | --- | --- | --- |
| children | adolescents | adults |  | No vaccination |  | Adults |  | Adults and adolescents |  |
| 1 | 0 | 1 | child | 0.25 | (0.13-0.40) | 0.029 | (0.014-0.050) | 0.029 | (0.014-0.050) |
| 1 | 0 | 1 | adult | 0.12 | (0.057-0.19) | 0.12 | (0.057-0.19) | NA |  |
| 0 | 1 | 1 | adolescent | 0.10 | (0.011-0.30) | 0.011 | (0.001-0.035) | 0.011 | (0.001-0.035) |
| 0 | 1 | 1 | adult | 0.12 | (0.057-0.19) | 0.12 | (0.057-0.19) | 0.12 | (0.057-0.19) |
| 2 | 0 | 2 | child | 0.48 | (0.33-0.62) | 0.24 | (0.16-0.31) | NA |  |
| 2 | 0 | 2 | adult | 0.20 | (0.097-0.31) | 0.13 | (0.065-0.21) | NA |  |
| 0 | 2 | 2 | adolescent | 0.13 | (0.014-0.37) | 0.043 | (0.0048-0.13) | 0.011 | (0.0012-0.036) |
| 0 | 2 | 2 | adult | 0.14 | (0.068-0.24) | 0.093 | (0.045-0.15) | 0.013 | (0.0060-0.022) |
| 2 | 2 | 2 | child | 0.50 | (0.32-0.67) | 0.31 | (0.21-0.42) | 0.16 | (0.11-0.21) |
| 2 | 2 | 2 | adolescent | 0.21 | (0.024-0.54) | 0.11 | (0.012-0.31) | 0.078 | (0.0086-0.20) |
| 2 | 2 | 2 | adult | 0.24 | (0.12-0.38) | 0.18 | (0.092-0.28) | 0.089 | (0.045-0.14) |

For the estimated parameters, vaccination with an effective but slightly leaky vaccine has the potential to strongly reduce the size of the household outbreaks (Table 2). For instance, in households consisting of a single child and a single adult, the SAR is 0.25 if the child is the primary case and no vaccination is applied, but just 0.029 if the adult has been vaccinated. In larger households, sizeable reductions can also be achieved, depending on the primary case. Focusing again on households of six persons, the SAR is reduced from 0.50 to 0.31 if a child is the primary case, from 0.21 to 0.11 if the primary case is an adolescent, and from 0.24 to 0.18 if the primary case is an adult. Adding vaccination of adolescents does further decrease household outbreak size in households in which an adolescent is present. However, due to the smaller rates of transmission to and from adolescents (Figure 3) the added benefit of adolescent vaccination is smaller compared to adult vaccination.

Second, we explore how adding adolescent vaccination might further reduce the probability of infection of a specific adult. This is especially relevant as adults have an intrinsically higher risk of severe disease, especially if the adult is immunocompromised or immunosuppressed [24, 25]. We focus on a number of illustrative scenarios for various household compositions and vaccination scenarios. The analyses show that the estimated probability of infection of the adult is high in the absence of vaccination, especially if a child is the primary infection in the household (Figure 4A), but can be strongly reduced by adult vaccination (Figure 4B). However, adding adolescent vaccination does not noticeably reduce the probability of adult infection further, as transmission is already strongly reduced and adults are already protected directly by vaccination (Figure 4C).

**Figure 4:**
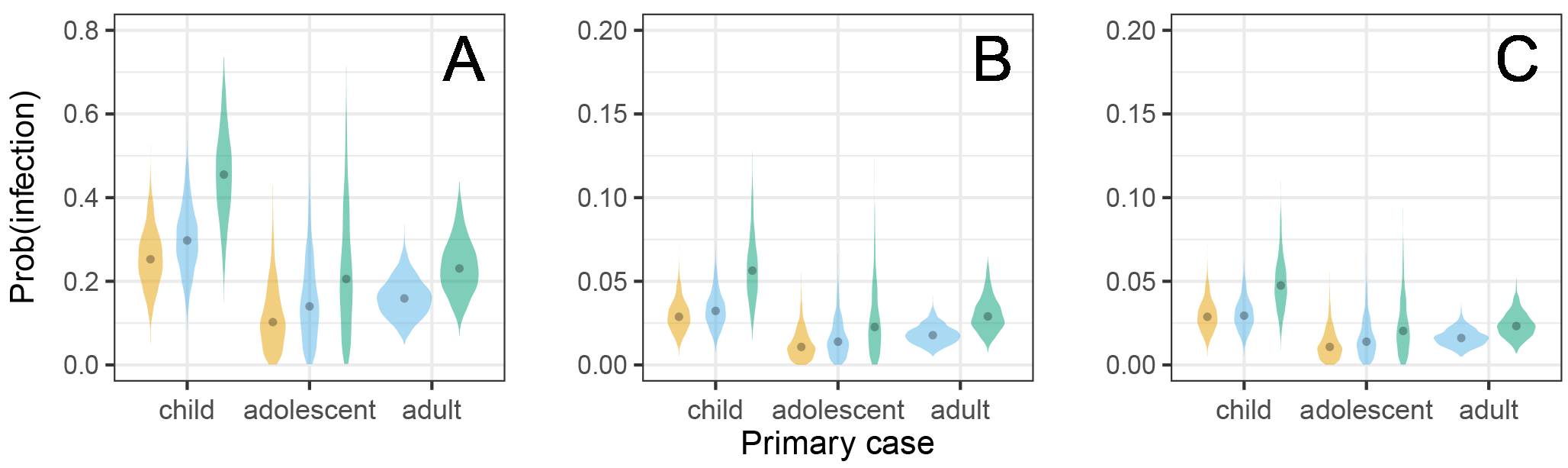
Estimates of the probability of infection of an adult for different household compositions, primary infection (child, adolescent, adult), and vaccination strategies. (A) no vaccination, (B) vaccination of adults, and (c) vaccination of adults and adolescents (C). Plots represent the posterior distribution (1,000 samples), and black dots indicate posterior medians. Vaccine efficacy for susceptibility is VE_*S*_ = 0.9. Notice the difference in scale on the y-axis between (A) and (B)-(C).

## Discussion

We have shown that precise estimates of SARS-CoV-2 household introduction hazards as well as within-household transmission rates can be obtained in a modestly sized study. This has been possible by virtue of the prospective set-up in which households are included before the first infection in the household has been observed. The prospective design has the added benefit compared to reactive household studies that there is lower risk of bias, in particular bias caused by preferential inclusion of larger households (as it is more likely that an infection is introduced in a larger than in a smaller household) and by preferential inclusion of households with severe infections (as these are more likely noticed) [4,26]. To make this design work logistically, we have employed a so-called passive-active follow-up strategy in which household are semi-passively followed during the at-risk period with an app, and in which more intensive follow-up is performed upon notification of an acute respiratory symptoms in the household [3].

With regard to the introduction hazards we have taken an agnostic approach in which hazards are optimally informed by the data, i.e. without assuming a specific underlying population-level transmission model. This was done on purpose, as the early SARS-CoV-2 pandemic has been strongly affected by lock-downs and behavioral responses, and is not easily described by simple transmission models [27–29]. With regard to within-household transmission, however, we fit a stochastic transmission model to the data. The within-household analyses provide estimates of age-stratified transmission rates, and in particular yield estimates of intrinsic transmissibility between children, adolescents, and adults in a population with low vaccination coverage and low pre-existing cumulative infection attack rates (*≈* 10 − 20%, [30]). Our analyses have revealed that, compared with adolescents and adults, children were not the main source of introduction of SARS-CoV-2 into households, that adolescents played a minor role propagating the infection in the household, that children were the dominant transmission source in the household (see also [2, 18, 26]), and in general that quantitative estimates of introduction hazards can be obtained. In fact, we are not aware of other studies providing such estimates that are optimally informed by the data. In our analyses, the estimated introduction hazards range from *≈* 0.0001 per child per day in epidemic troughs to *≈* 0.001 per adolescent/adult per day in epidemic upswings. Timing of the peaks and troughs relative to hospital admissions in the Netherlands (Figure 2) suggest that this approach has produced reliable results.

While we are convinced that within the confines of our data we have selected a transmission model that is in a statistical sense optimal, we also acknowledge a number of weaknesses and alternatives. First, throughout we have assumed a so-called density-dependent transmission model in which each individual in the household makes a fixed number of contacts with each of the other household members per unit of time. This was done for convenience, as it enables direct translation of the estimated transmission parameters to conditional infection probabilities irrespective of household size, but also because this model provides a slightly better fit to the data than a frequency-dependent transmission model. However, our data contains just 59 infected households, and has limited variation in the size of infected households (3-6 persons). Therefore, alternative contact scenarios cannot yet be discarded (see also [22]). Reassuringly, estimates of person-to-person transmission probabilities are very close under the density- and frequency-dependent contact models for the most common household composition of four persons.

Second, we have assumed that if initial cases in the household are found at the same day, that these cases represent co-primary cases. If one or more of these co-primary cases would actually have been infected in the household, then including this information in the analyses increases within-household transmission rates while decreasing introduction hazards (see Results). In general, it will be very difficult to determine which person or persons have been the primary case or primary cases if onsets of symptoms are on the same day or only a few days apart. This inability to pinpoint the primary case is common to all household-based studies, and is a weakness that is not easily remedied. Fortunately, in sensitivity analyses our results appeared qualitatively robust to such trade-offs between introduction and transmission rates when more or fewer infections are assumed to represent primary case(s).

A third point of concern is that temporal information is used to estimate the introduction hazard but that only limited temporal information is used in the within-household analyses. In fact, temporal information only affects the probability that an additional infection will be introduced from outside the household. We have assumed that household outbreaks have a duration that is equal to the outbreak durations as defined earlier [3]. In these analyses most household outbreaks had a duration of 2 to 5 weeks (median: 26 days, range 13-126 days). Fortunately, the parameters seemed to be hardly affected by assumptions on the duration of the household outbreaks, as the estimated introduction hazard is very small compared to the within-household transmission rates (¿2 orders difference). For instance, setting the outbreak duration to either 14 or 28 days for all households hardly affected the results. More problematic, though, might be the implicit assumption that the introduction hazard may vary over time and by person-type, but does not depend on whether other household members have recently been infected outside the household. This can be unrealistic if household members share contacts outside of the household. Incorporation of such correlations in the introduction hazard is a major direction of future model development and applications.

Our findings demonstrate that prospective household-based studies hold considerable promise to study the interplay between vaccination- and infection-induced immunity on the household introduction hazard, the susceptibility to (re-)infection, and the infectiousness once infected in terms of biologically interpretable parameters. In addition, analysing such data using transmission models has the added advantage over traditional statistical analyses [2,26,31] that transmission model-based estimates can feed into impact analyses of interventions.

## Methods

### Study design and data collection

The Cokids household-based study into the causes of acute respiratory infections (ARIs) in households with underage children had been conducted between August 2020 to August 2021. Enrollment was focused on the period between August 2020 and February 2021. Eligible households had been selected from three existing Dutch birth cohort studies, and all had at least one child aged 0-17 years. Details of the study design are presented elsewhere [3]. Here we briefly mention the salient aspects of the study relevant to our analyses. First, the study contained a core study with follow-up of a maximum of 161 days, and an extended study with follow-up period until July 2021. Since follow-up criteria are different between the core and extended follow-up, and could potentially lead to bias, we here only include the basic follow-up period for estimation of the introduction hazards. We did, however, include 4 households in the extended follow-up in the analyses of within-household transmission. Additionally, there were 4 households of size 2 in the core study that provided information on the introduction of SARS-CoV-2 into the household but not on within-household transmission as both persons were labelled as primary cases. There was only 1 vaccinated person in the data used for our analyses.

All participants were checked daily, using an app developed specifically for the study, for the occurrence of respiratory symptoms and fever. In addition, all participants were screened for a panel of respiratory viruses at a 4 to 6 weeks time interval, irrespective of symptoms. An outbreak study with intensified follow-up of the household was initiated when (1) a household member developed new-onset respiratory symptoms or fever, or (2) a SARS-CoV-2 positive result was received on a screening test, or (3) a positive test result was received from an external testing site. Details of the follow-up procedures are given elsewhere [3].

Hospital admission data have been retrieved from the National Institute for Public Health and the Environment’s open data (https://data.rivm.nl/covid-19). To remove weekday effects we present these data as a centered 7-day moving average.

### Transmission model

Our analyses are based on a multi-type susceptible-exposed-infectious-recovered (SEIR) transmission model. In this model individuals are classified as susceptible to infection (S), infected but not yet infectious (E), infected and infectious (I), or recovered and immune (R). Throughout we stratify the population by age as follows: children (under 12 years), adolescent (12-18 years), and adults (18 years and older). This stratification corresponds well with age at which children transition from primary to secondary school, and from secondary school to subsequent education in the Netherlands [2, 3].

The within-household analyses use the number of infections that have occurred at the end of the household outbreaks [8, 11,23, 32]. Statistical methodology based on such final size data is well-developed, and these methods have advantages over analyses that use all temporal information. First, the number of infections in the household can usually be determined with high certainty, while the timing of transitions between classes is often uncertain. Second, final size analyses are invariant with respect to the latent period, i.e. the time that individual spend in the exposed (E) class [32]. Hence we may, without loss of generality, focus on a simplified SIR type model with no latent period. Third, final size data do not allow estimation of parameters with respect to calendar time, but only relative to other model parameters. This enables simplification of the model by reducing the number of parameters. Specifically, we can assume, again without loss of generality, that the mean of the infectious period is fixed at length 1 time unit, and set the basic reproduction number equal to the contact rate parameter in a reference class. Here, we assume that adults are the reference class. Variation in the infectious periods can affect the outcomes, however, and we assume that the infectious period is gamma distributed such that 95% equal-tailed coverage of the infectious period ranges from 0.74 to 1.30, i.e. approximately 6-10 days when the mean infectious period is 8 days [33]. We supplement the default analyses with a scenario in which the infectious period is fixed at 1 time unit (i.e. delta-distributed), and with a scenario with increased variation in the infectious period such that the 95% coverage of the infectious period distribution ranges from 0.61 to 1.48 of the mean.

Further, with respect to the number of contacts in households of different sizes we focus on two extremes, viz a density-dependent contact model in our default scenario, and a frequency-dependent contact model as alternative [34]. In the density-dependent model each household member makes an identical expected number of contacts *with each of the other household members* per unit of time, while in the frequency-dependent transmission model each household member makes an identical number of contacts per unit of time. Hence, in the frequency-dependent model fewer contact are made with each of the household members in a larger than in a smaller household.

### The final size distribution

The statistical methods rely on the fact that we can compute the probability distribution of the final outbreak size of a given household. Let *a, n* and 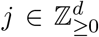 represent the number of primary household cases, initially uninfected members and the number of secondary infections, respectively. Here *d* is the number of types (in our analysis *d* = 3 age classes). Given fixed *a* and *n*, we want to calculate the probabilities *Q*_*j*_ of the final size 0 *≤ j ≤ n*. The probabilities *Q*_*j*_ depend on a number of parameters. First, we require the transmission rate matrix 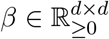, in which the element *β*_*μν*_ denotes the transmission rate from an individual of type *ν* to type *μ*. Next, let *b* denote the vector of probabilities of escape from external infection, i.e., 1 *− b*_*μ*_ is the probability that an individual of type *μ* gets infected during the household outbreak by someone from outside the household. Finally, the duration of each infection is an i.i.d. random variable *T*_*i*_. The probabilities *Q*_*j*_ are expressed in terms of the Laplace transform of *T*_*i*_, given by 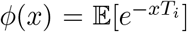. In our analysis *T*_*i*_ *∼* Gamma(*α, α*^−1^), such that 𝔼 [*T*_*i*_] = 1 time unit, and *ϕ*(*x*) = (1 *− x/α*)^*−α*^. For integer vectors *m* and *ℓ* and a real vector *x*, we write 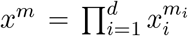 and 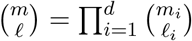. For the scalar function *ϕ*, we write *ϕ*(*x*) to denote the vector (*ϕ*(*x*_1_), …, *ϕ*(*x*_*d*_))^*′*^. With these definitions and notation in place, the final size probability *Q*_*j*_ is given by 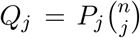 where *P*_*j*_ satisfies the recursive equation

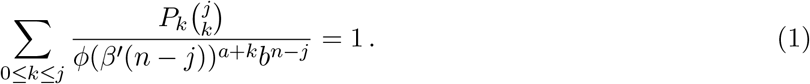

Although formal derivations of Eq (1) can be found elsewhere [23, 32], here we give an intuitive derivation using elementary methods. In particular, our analyses do not make use of a Wald identity. We present the method for *d* = 1 (i.e. no age stratification), but the arguments easily generalize to *d >* 1.

We write *m* : *ℓ* to denote the set of indices *m, m* + 1, …, *ℓ*, which is empty when *m > ℓ*. First, we enumerate the non-primary members of the household as 1 : *n*, and we split them into two groups: 1 : *j* and *j* + 1 : *n*. If we condition on how many members in the first group get infected, then we can easily calculate the probability that none of the second group get infected. Suppose that *k* individuals of 1 : *j* get infected, then this conditional probability equals

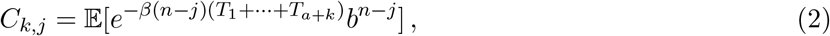

where *T*_1_, …, *T*_*a*+*k*_ are the gamma-distributed lengths of the infectious periods of the *a* primary cases and *k* infected members in group 1 : *j*. The infectious periods of infected members can overlap, in which case we assume the transmission hazard is additive. With probability *b*^*n−j*^, no one in group *j* + 1 : *n* gets infected due to external contacts. The lengths *T*_*i*_ are unknown, and therefore we take the expectation to integrate them out. As the *T*_*i*_ are i.i.d., we get *C*_*k,j*_ = *ϕ*(*β*(*n − j*))^*a*+*k*^*b*^*n−j*^. Next, let *P*_*k*_ denote the probability that exactly individuals 1 : *k* are infected. If *k ≤ j*, then we can interpret *P*_*k*_ as the probability that 1 : *k* of the first group 1 : *j* are infected, and none of *j* + 1 : *n* are infected. Moreover, the product 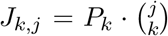 is equal to the joint probability that *k* arbitrary members of group 1 : *j* are infected (as opposed to exactly members in 1 : *k*), and none of *j* + 1 : *n*. Finally, let *U*_*k,j*_ denote the unconditional probability that *k* members of 1 : *j* are infected (regardless of what happens to *j* + 1 : *n*), then we find using the definition of conditional probability that *P*_*k*_, and *U*_*k,j*_ are related by

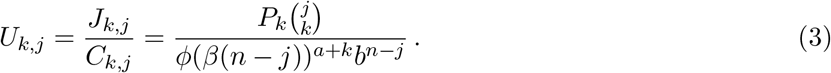

Using the law of total probability, we find that 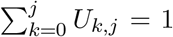, and hence we find the recursive Eq (1) for *P*_*k*_, which includes the edge case *P*_0_ = *ϕ*(*βn*)^*a*^*b*^*n*^, and therefore allows us to compute *P*_*j*_. Since the household division 1:*j* and *j*+1:*n* was arbitrary, we can now compute the final size probability 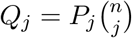.

### Hazard of external infection

To estimate the hazard of external infection we use semi-parametric penalized splines [35]. A related but simpler approach in which a fixed introduction probabilities are estimated for predefined periods is given in House *et al* [20].

In our framework, the spline *p*(*t*) is defined as a linear combination of cubic basis splines 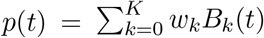, with 50 equidistant knots such that *K* = 52. To penalize large deviations of *p*(*t*), the weights *w* are equipped with a random-walk prior as follows. We choose *w*_0_ ∼ 𝒩 (−7.5, 2.5) and define the other weights cumulatively as 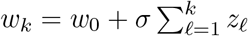, where *z*_*ℓ*_ *∼* 𝒩 (0, 10). The diffusion parameter *s* determines the smoothness of the spline and is given a weakly-informative prior *s*^2^ *∼* InvGamma(1, 0.0005). For the adult age class, we then define the hazard of infection *h*_3_(*t*) = exp(*p*(*t*)). The hazards for the other age classes are proportional to the adult hazard, i.e. *h*_1_(*t*) = *r*_1_*h*_3_(*t*) for children and *h*_2_(*t*) = *r*_2_*h*_2_(*t*) for adolescents, where *r*_1_, *r*_2_ > 0 are the relative hazards (Table 2).

### Likelihood function

With the ingredients specified above, we can formulate the likelihood *L*(*t, a, n, j*) of observing a household infected at time *t* with *a* primary infected persons, *n* non-primary persons, and *j* persons infected over the course of the household outbreak:

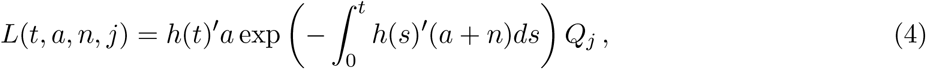

which is the product of the likelihood that the index cases are infected at time *t*, and the probability of final size *j*. This probability *Q*_*j*_ depends on *n* and *a*, but also on *t*, because the probability of escape *b* depends on the external infection hazard *h* as follows: 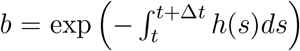, where Δ*t* is either set at 14 or 28 (days), or is defined as the household-specific period that an infected household is monitored. For households that were not infected during the study period, the final size is unobserved and the introduction time is right-censored. Therefore, the likelihood is given by

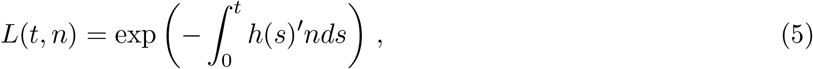

where *t* is time that the household dropped out of the study and *n* is the household composition. In our inferential analyses, we approximate the cumulative hazards with sums 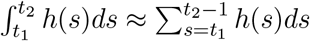.

### Inference and model selection

We estimate the parameters in a Bayesian framework using Hamiltonian Monte Carlo. Except for the weakly-informative weights of the penalized spline (see above), none of the other parameters are given explicit prior distributions. Hence, these parameters have (improper) uniform prior distributions on their domains, making each possible parameter value equally likely a priori. To reduce correlations between parameters and facilitate mixing, we parameterize the proportionate mixing model and simplifications thereof in terms of absolute infectivities *f*_*i*_ and absolute susceptibilities *g*_*i*_ (*i* = 1, 2, 3), such that the transmission rates are given by *beta*_*i,j*_ = *g*_*i*_*f*_*j*_, and in particular the transmission rate in the reference class (*i* = 3, adults) is given by *β* ≡ *β*_3,3_ = *g*_3_*f*_3_. Using this formulation the relative infectivities and susceptibilities of the other person-types (Table 2) are given by 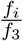 and 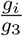 *i* = 1, 2). Since one parameter is redundant, we have further taken *g*_3_ ≡ 1 without loss of generality.

All analyses are performed using Stan (version 2.29.0) and R (version 4.2.2) using the CmdStanR package (version 0.4.0) as interface between R and Stan [36]. We run 10 MCMC chains in parallel and base the analyses on 1,000 samples from 10 chains, where we have applied 1/10 thinning. In all model runs effective number of samples generally ranges from 900-1,100, while the convergence criterion 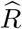 ranges from 0.99-1.01. Data, scripts, and figures are available in the online repository at https://github.com/mvboven/sars2-households.

Model selection is based on information criteria for singular statistical models [37, 38]. Specifically, we use LOO_IC from the loo package (version 2.4.1) to gauge relative predictive performance, and calculate WBIC using a run of the model at the appropriate sampling temperature to select the most likely data generating process. As estimation of the introduction hazards is already optimized for predictive performance, we have used LOOIC and WBIC only for the within-household analyses in model runs that exclude external infection during the household outbreaks.

## Data Availability

All data used in this study are available at https://github.com/mvboven/sars2-households.

## Acknowledgments and funding

We gratefully acknowledge all participants of the study. This study was funded by the Netherlands Organization for Health Research and Development (ZonMw) (grant 10150062010006) and the National Institutes of Health (NIH) (grants P01-AI131365 and R01-OD011095).

